# Optimal combinations of CSF biomarkers for predicting cognitive decline and clinical conversion in cognitively unimpaired participants and mild cognitive impairment patients: A multi-cohort study

**DOI:** 10.1101/2022.09.07.22279676

**Authors:** Gemma Salvadó, Victoria Larsson, Karly A Cody, Nicholas C Cullen, Erin M Jonaitis, Erik Stomrud, Gwendlyn Kollmorgen, Norbert Wild, Sebastian Palmqvist, Shorena Janelidze, Niklas Mattsson-Carlgren, Henrik Zetterberg, Kaj Blennow, Sterling C Johnson, Rik Ossenkoppele, Oskar Hansson

**Affiliations:** Clinical Memory Research Unit, Department of Clinical Sciences Malmö, Lund University, Lund, Sweden; Wisconsin Alzheimer’s Disease Research Center University of Wisconsin School of Medicine and Public Health Madison Wisconsin, Madison, Wisconsin, USA; Wisconsin Alzheimer’s Institute, School of Medicine and Public Health, University of Wisconsin-Madison, Madison, Wisconsin, USA; Memory Clinic, Skåne University Hospital, Malmö, Sweden; Roche Diagnostics GmbH, Penzberg, Germany; Wallenberg Center for Molecular Medicine, Lund University, Lund, Sweden; Department of Neurology, Skåne University Hospital, Lund, Sweden; Department of Psychiatry and Neurochemistry, the Sahlgrenska Academy at the University of Gothenburg, Mölndal, Sweden; Clinical Neurochemistry Laboratory, Sahlgrenska University Hospital, Mölndal, Sweden; Department of Neurodegenerative Disease, UCL Institute of Neurology, Queen Square, London, United Kingdom; UK Dementia Research Institute at UCL, London, United Kingdom; Hong Kong Center for Neurodegenerative Diseases, Hong Kong, China; Department of Medicine, School of Medicine and Public Health, University of Wisconsin-Madison, Madison, Wisconsin, USA; Geriatric Research, Education and Clinical Center at the William S. Middleton Memorial Veterans Hospital, Madison, Wisconsin, USA; Alzheimer Center Amsterdam, Department of Neurology, Amsterdam Neuroscience, Vrije Universiteit Amsterdam, Amsterdam University Medical Center, Amsterdam, the Netherlands

**Author notes:** **Corresponding author information:** Oskar Hansson, MD, PhD, Memory Clinic, Skåne University Hospital, SE-205 02, Malmö, Sweden. These authors contributed equally as first authors (Gemma Salvadó, Victoria Larsson).

**Keywords:** Cognitive decline, conversion to dementia, phosphorylated tau, amyloid-β, neurodegeneration, glial activation, inflammation, BioFINDER, WRAP, WADRC

## Abstract

**INTRODUCTION:** Our objective was determining the optimal combinations of cerebrospinal fluid (CSF) biomarkers for predicting disease progression in Alzheimer’s disease (AD) and other neurodegenerative diseases.

**METHODS:** We included 1,983 participants from three different cohorts with longitudinal cognitive and clinical data, and baseline CSF levels of Aβ42, Aβ40, p-tau, NfL, neurogranin, α-synuclein, sTREM2, GFAP, YKL-40, S100b and IL-6 (Elecsys® NeuroToolKit).

**RESULTS:** Change of modified Preclinical Alzheimer’s Cognitive Composite (mPACC) in cognitively unimpaired (CU) was best predicted by p-tau/Aβ42 alone (R^2^≥0.31) or together with neurofilament light ([NfL],R^2^=0.25), while p-tau/Aβ42 (R^2^≥0.19) was sufficient to accurately predict change of the Mini-Mental State Examination (MMSE) in mild cognitive impairment (MCI) patients. P-tau/Aβ42 (AUC≥0.87) and p-tau/Aβ42 with NfL (AUC≥0.75) were the best predictors of conversion to AD and all-cause dementia, respectively.

**DISCUSSION:** P-tau/Aβ42 is sufficient for predicting progression in AD, with very high accuracy. Adding NfL improves the prediction of all-cause dementia conversion and cognitive decline.

## Background

Amyloid-β (Aβ) plaques and neurofibrillary tau tangles are key pathological hallmarks in Alzheimer’s disease (AD) [1]. Cerebrospinal fluid (CSF) measures of these two pathologies have shown high accuracy for the diagnosis and prognosis of AD [2,3] and are currently, together with positron emission tomography (PET), used for diagnosis in the clinical practice. CSF biomarker development has also enabled measurement of other pathophysiological alterations related to AD and other dementias. Some of these biomarkers may provide additional information on individuals’ disease stage and progression, while others may enhance the understanding of underlying biological processes occurring during the course of the disease. Some of the most promising novel CSF biomarkers are related to neurodegeneration and microglial or astroglial activation [4] (see [5] and [6] for extensive reviews). However, until recently, these biomarkers had to be measured in different platforms and using sophisticated methods, which reduced its applicability to clinical practice.

Some of these biomarkers have recently been implemented in a single panel of automated CSF immunoassays, with the Elecsys® NeuroToolKit platform (Roche Diagnostics International Ltd, Rotkreuz, Switzerland), facilitating implementation and direct comparison between different centres. This panel measures biomarkers of neurodegeneration (neurofilament light [NfL], neurogranin and α-synuclein); glial activity and neuroinflammation (soluble triggering receptor expressed on myeloid cells 2 [sTREM2], glial fibrillary acidic protein [GFAP], YKL-40 [also known as chitinase 3-protein 1], S100 calcium-binding protein B [S100b] and interleukin 6 [IL-6]); as well as core AD-biomarkers (phosphorylated tau at threonine-181 [p-tau], Aβ42 and Aβ40). Previous studies using this panel have shown alterations in some of these biomarkers in different stages and in relation to different aspects of the disease [7–10]. However, their clinical utility for predicting disease progression after accounting for core AD-biomarkers is still largely unknown. Studies in large cohorts from different centres are also needed to verify generalizability of findings related to these biomarkers.

Therefore, we aimed to identify the best combination of CSF biomarkers for an accurate prediction of cognitive decline and clinical conversion, using longitudinal data from three large cohorts. More specifically, we aimed to assess whether the core-AD biomarkers (operationalized as the CSF p-tau/Aβ42 ratio and the CSF Aβ42/40 ratio) would be sufficient to accurately predict disease progression or whether biomarkers targeting other pathophysiological mechanisms during AD could significantly improve their prediction.

## Methods

### Participants

Participants were included from three cohorts: the Swedish BioFINDER-1 (NCT01208675) [11] and BioFINDER-2 (NCT03174938) [12] at Lund University (Lund, Sweden), and a joint cohort comprised of subsets who met inclusion criteria from Wisconsin Registry for Alzheimer’s Prevention (WRAP) and Wisconsin Alzheimer’s Disease Research Center (WADRC) at Wisconsin University (Wisconsin, USA) [13]. In both BioFINDER-1 and BioFINDER-2 cognitively unimpaired (CU) people were recruited from population-based studies in the city of Malmö as previously described [11,12]. These individuals did not fulfil the criteria of mild cognitive impairment (MCI) or dementia at baseline. Further, The BioFINDER studies included patients with either subjective cognitive decline (SCD) or MCI (none of one fulfilled the criteria for any type of dementia at baseline) from the Memory clinics in southern Sweden. Following the NIA-AA recommendations cognitive normal and subjects with SCD were defined as cognitively unimpaired [14]. Exclusion criteria included 1) significant unstable systemic illness or organ failure, such as terminal cancer, that makes it difficult to participate in the study. 2) Current significant alcohol or substance misuse, or 3) Refusing lumbar puncture or neuropsychological assessment. Participants from WRAP and WADRC include cognitively unimpaired and impaired participants, who are enriched for parental family history of AD, and undergo neuropsychological evaluations on an annual or biennial basis. More information about the recruitment of these participants can be found in [13], but in summary, participants were included from memory clinics in which a parent was diagnosed or treated, advertisements, and word of mouth. The participants of the present study were enrolled between November 2007 to May 2020. All participants gave written informed consent and ethical approval was granted by the Regional Ethical Committee in Lund, Sweden and University of Wisconsin Health Sciences Institutional Review Board, respectively.

### CSF measurements

CSF levels of Aβ42 [15] as well as Total-Tau and Phospho-Tau(181P) [16] were measured using the Elecsys β-Amyloid (1–42), Total-Tau and Phospho-Tau(181P) CSF electrochemiluminescence immunoassays on a fully automated cobas e 601 instrument (Roche Diagnostics International Ltd., Rotkreuz, Switzerland). The other CSF biomarkers (Aβ40, NfL, neurogranin, YKL-40, GFAP, sTREM2, S100b, IL-6 and α-synuclein) were measured with robust prototype assays as part of the Roche NeuroToolKit on cobas e 411 and e 601 instruments (Roche Diagnostics International Ltd, Rotkreuz, Switzerland). All measurements were performed at the Clinical Neurochemistry Laboratory, University of Gothenburg, Sweden by board-certified laboratory technicians who were blinded to diagnostic and other clinical data. We also calculated the CSF p-tau/Aβ42 and the Aβ42/Aβ40 ratios. These ratios were included in this study as they may represent reliable alternatives for amyloid PET [17–21] and are often used in the clinical setting for diagnostic [22–26] and/or prognostic purposes [27,28], which is the main objective of this study. However, we acknowledge that these may not be a direct measure of AD-related brain pathology.

### Outcomes

The primary cognitive outcome for CU was the modified Preclinical Alzheimer’s Cognitive Composite (mPACC) given its previously proven sensitivity to detect changes in cognition in CU participants [29]. The mPACC in BioFINDER-1 and BioFINDER-2 was calculated as the average of four z-scores, for tests of memory (the delayed recall test from the cognitive subscale from the Alzheimer’s Disease Assessment Scale [ADAS-cog]) with a double weight to preserve the weight of memory tests in the original PACC [29], verbal ability (animal fluency), executive function (Trail Making Test B [TMT-B]), and global cognition using the Mini-Mental Estate Examination (MMSE), as previously described [30]. In WRAP and WADRC mPACC was calculated as the mean of z-scores of three tests: TMT-B for timed executive function, delayed story recall harmonized from either the Craft Story (for WADRC) or WMS-R Logical Memory story A (for WRAP), and the total over trials of the Rey Auditory-Verbal Learning Test (AVLT) [31]. For MCI patients, cognitive decline was measured with MMSE in all cohorts. In the joint Wisconsin cohort some MMSE scores were obtained through conversion from Montreal Cognitive assessment (MOCA) scores as previously described [32].

### Statistical analyses

Two main analyses were performed aiming at identifying the best combination of CSF biomarkers to predict either cognitive decline or clinical conversion. The biomarkers included as individual predictors in all the analyses were: the p-tau/Aβ42 ratio, the Aβ42/40 ratio, NfL, neurogranin, YKL-40, GFAP, sTREM2, S100b, IL-6 and α-synuclein. To study cognitive decline, we used mPACC for CU individuals and MMSE for MCI participants, independently for each cohort. Linear mixed models were used to predict cognitive decline using random intercept and random time slopes with baseline age, sex, *APOE-ε4* carriership and education as covariates. The model including only covariates and time was considered the basic model. To select the best model including baseline CSF biomarkers, a forward selection procedure was used based on Bayesian information criteria (BIC) using the *aba* package for R (v.4.1.0). For all biomarkers included in the model, we also included their interaction with time. For each case (*i*.*e*., cognitive measure and cohort), a parsimonious model (*i*.*e*., best prediction with the lowest number of predictors) was selected with a forward step-wise inclusion of the most significant biomarkers until the difference in BIC with the next model (*i*.*e*., including an additional biomarker) was not significant (ΔBIC>6), which indicates strong evidence of improvement [33]. When different, the parsimonious models were compared with those only including covariates and CSF p-tau/Aβ42 ratio, as a typical measure of AD pathology used in the clinical setting [2].

For predicting clinical conversion, we followed a similar approach using generalized linear models including a binomial family. Progression to AD dementia and to all-cause dementia were studied independently in each cohort for both CU and MCI. Models with less than 20 cases converting to dementia were disregarded. Basic models included age, sex, and *APOE-ε4* carriership as covariates. Best models were selected also using a forward selection procedure with the *aba* package based on differences in area under the curve (AUC). Models were selected as preferred based on AUC using the DeLong’s test to check for significant differences (p<0.05). Best models were also compared with those only including covariates and CSF p-tau/Aβ42 ratio. We also computed hazard ratios (HR) for each biomarker selected in each of the parsimonious models using cox proportional hazards regression models, and constructed Kaplan-Meier curves using the *survival* package from R. Finally, we repeated all the main analyses with data truncated at 4- and 6-years of follow-up

## Results

### Participants

A total of 1,453 CU and 530 MCI patients were included in this study. The mean (SD) age of CU individuals was 66.6 (9.7) years, 857 (59.0%) of them were women and 553 (38.1%) were *APOE-ε4* carriers. For MCI patients, the mean (SD) age was 71.2 (7.2) years, and there were 225 (42.5%) women and 269 (50.8%) *APOE-ε4* carriers. CU were followed for a mean (SD) time of 4.7 (3.1) years and MCI for 2.8 (2.2) years. Baseline characteristics are presented in Table 1. Out of the information available, 43/1,000 (4.3%) CU participants converted to AD dementia and 66/1,000 (6.6%) to all-cause dementia. From MCI at baseline patients, 185/529 (35.0%) converted to Alzheimer’s dementia and 297/529 (56.1%) to all-cause dementia.

**Table 1.** Participant Characteristics. * Cognitive measure was mPACC for CU and MMSE for MCI patients. Abbreviations: CU, cognitively unimpaired; MCI, mild cognitive impairment; MMSE, Mini-Mental State Examination; mPACC, modified Preclinical Alzheimer Cognitive Composite; WADRC, Wisconsin Alzheimer’s disease Research Center; WRAP, Wisconsin Registry for Alzheimer’s Prevention.

|  | **BioFINDER-1** |  | **BioFINDER-2** |  | **WRAP & WADRC** |  |
| --- | --- | --- | --- | --- | --- | --- |
|  | CU<br>(n=550) | MCI<br>(n=260) | CU<br>(n=453) | MCI<br>(n=215) | CU<br>(n=450) | MCI<br>(n=55) |
| Age, mean (SD)<br>[range] | 71.8 (6.09)<br>[41.3, 88.4] | 71.5 (5.44)<br>[60.1, 80.8] | 65.7 (11.6)<br>[40.5, 88.7] | 70.9 (8.37)<br>[43.1, 93.3] | 61.2 (7.64)<br>[42.7, 85.5] | 70.6 (9.07)<br>[54.1, 87.3] |
| Women, No (%) | 312 (56.7%) | 107 (41.2%) | 242 (53.4%) | 98 (45.6%) | 303 (67.3%) | 20 (36.4%) |
| *APOE-ε4* carriers,<br>No (%) | 181 (32.9%) | 129 (49.6%) | 204 (45.0%) | 113 (52.6%) | 168 (37.3%) | 27 (49.1%) |
| Years of<br>education, mean<br>(SD) | 12.4 (3.6) | 11.0 (3.3) | 12.6 (3.4) | 12.5 (4.1) | 16.2 (2.5) | 15.9 (2.7) |
| Years of follow-up, mean (SD) | 6.2 (2.2)<br>[0, 10.9] | 3.3 (2.1)<br>[0.35, 8.5] | 1.2 (1.04)<br>[0, 3.6] | 1.5 (1.10)<br>[0, 3.6] | 6.6 (2.34)<br>[0.9, 11.2] | 5.2 (2.47)<br>[1.1, 10.0] |
| Cognitive<br>measure, mean<br>(SD)* | 0.041<br>(0.719) | 27.0<br>(1.9) | 0.028<br>(0.764) | 27.0<br>(1.9) | 0.108<br>(0.704) | 27.6 (2.1) |
| Missing, No (%) | 96 (17.5%) | - | 5 (1.1%) | 1 (0.5%) | 33 (7.3%) | 1 (1.8%) |
| Conversion to<br>Alzheimer's-type<br>dementia, No (%) | 39 (7.1%) | 117 (45.0%) | - | 50 (23.3%) | 4 (0.9%) | 18 (32.7%) |
| Missing, No (%) | - | 1 (0.4%) | - | - | - | - |
| Conversion to all-cause dementia,<br>No (%) | 61 (11.1%) | 189 (72.7%) | - | 84 (39.1%) | 5 (1.1%) | 24 (43.6%) |
| Missing, No (%) | - | 1 (0.4%) | - | - | - | - |

### Cognitive decline

Parsimonious models for predicting cognitive decline can be found in Table 2. In summary, mPACC change was best predicted by CSF p-tau/Aβ42 ratio alone in BioFINDER-2 and WRAP & WADRC (BioFINDER-2: β[95%CI]= -0.10 [-0.14, - 0.05], R^2^=0.36; WRAP & WADRC: β[95%CI]= -0.12 [-0.15, -0.09], R^2^=0.31). In BioFINDER-1, the addition of CSF NfL (β[95%CI]= -0.09 [-0.12, -0.05]) to CSF p-tau/Aβ42 ratio (β[95%CI]= -0.15 [-0.19, -0.12]) significantly improved this prediction (with NfL: R^2^=0.25 vs. without NfL: R^2^=0.20, ΔBIC=26, p<0.001). The parsimonious model to predict change in MMSE in MCI patients only included CSF p-tau/Aβ42 ratio in all three cohorts (BioFINDER-1: β[95%CI]= -0.24 [-0.30, -0.17], R^2^=0.40; BioFINDER-2: β[95%CI]= -0.14 [-0.23, -0.05] R^2^=0.19; WRAP & WADRC: β[95%CI]= -0.32 [-0.49, -0.15], R^2^=0.32). However, in BioFINDER-2 this model was not significantly different than the basic model only including covariates. Depiction of cognitive decline and predictions using the parsimonious models are shown in Figure 1. Sensitivity analyses with cognitive data truncated at 4 and 6 years of follow-up can be found in Supplementary Tables 1 and 2, respectively.

**Table 2.** Parsimonious model description for predicting cognitive decline. Description of the parsimonious model for predicting cognitive decline for each diagnosis group and cohort. These models were derived independently per each cohort and baseline diagnosis. Only those biomarkers selected to be into the parsimonious model per cohort and disease stage are shown in each case. Effect sizes (β) for each selected biomarker and statistics (BIC and R^2^) of the final model are shown. BIC and R^2^ of the basic model (only covariates) are included for comparison. Linear mixed models with random slope and intercept were used in all cases. β estimates represent the effect size of each biomarker’s interaction with time. Cognitive decline was assessed with mPACC in CU and with MMSE in MCI patients. Covariates were included in all models and were: age, sex, *APOE-ε4* carriership, education and time. Abbreviations: Aβ, amyloid-β; BIC, Bayesian Information Criterion; CI, confidence interval; CU, cognitively unimpaired; MCI, mild cognitive impairment; MMSE, Mini-Mental State Examination; mPACC, modified Preclinical Alzheimer Cognitive Composite; p-tau, phosphorylated tau; WADRC, Wisconsin Alzheimer’s Disease Research Center; WRAP, Wisconsin Registry for Alzheimer’s Prevention.

| Diagnosis group | Cohort | Biomarker | $\beta$ [95% CI] | BIC parsimonious | $R^2$ parsimonious | BIC basic | $R^2$ basic |
| --- | --- | --- | --- | --- | --- | --- | --- |
| CU | BioFINDER-1 | p-tau/Aβ42 | -0.15<br>[-0.19, -0.12] | 3109.2 | 0.25 | 3230.6 | 0.06 |
|  |  | NfL | -0.09<br>[-0.12, -0.05] |  |  |  |  |
| | BioFINDER-2 | p-tau/Aβ42 | -0.10<br>[-0.14, -0.05] | 1297.9 | 0.36 | 1314.1 | 0.27 |
| | WRAP & WADRC | p-tau/Aβ42 | -0.12<br>[-0.15, -0.09] | 3631.6 | 0.31 | 3681.8 | 0.26 |
| MCI | BioFINDER-1 | p-tau/Aβ42 | -0.24<br>[-0.30, -0.17] | 2390.7 | 0.40 | 2426.9 | 0.30 |
| | BioFINDER-2 | p-tau/Aβ42 | -0.14<br>[-0.23, -0.05] | 976.6 | 0.19 | 978.8 | 0.13 |
| | WRAP & WADRC | p-tau/Aβ42 | -0.32<br>[-0.49, -0.15] | 582.0 | 0.32 | 594.79 | 0.14 |

**Figure 1.**
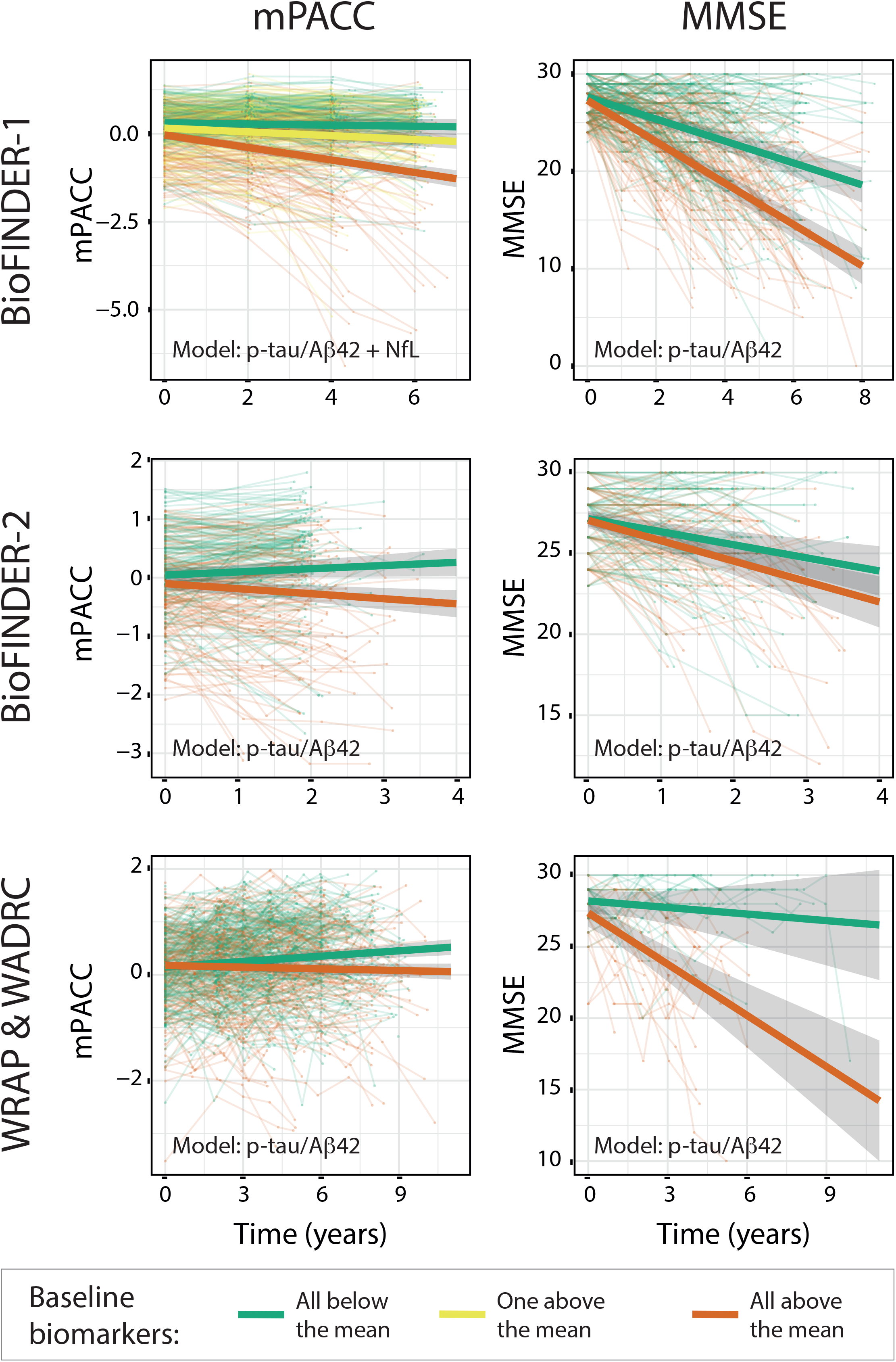
Cognitive change. Depiction of cognitive change over time per cohort and clinical group. Cognitive change in CU participants is shown with mPACC change (first column) while cognitive change in MCI patients is shown with MMSE (second column). Bold lines represent the predicted trajectory using the parsimonious model based on biomarkers levels at baseline. Green (red) lines represent participants with all biomarkers in the parsimonious model below (above) the mean at baseline. For the model with more than one biomarker included in the parsimonious model, the yellow line represents subjects with only one biomarker above the mean at baseline. Grey bands represent 95% confidence intervals. These lines are only for visualization purposes and represent the mean trajectory of the group of participants included in these artificial groups. Individual trajectories were calculated per each participant in the statistical model. Linear mixed models with random slope and intercept were used to construct the models. Biomarkers used in the parsimonious model in each case are detailed in the plots. Abbreviations: CU, cognitively unimpaired; MCI, mild cognitive impairment; MMSE, Mini-Mental State Examination; mPACC, modified Preclinical Alzheimer Cognitive Composite; WADRC, Wisconsin Alzheimer’s disease Research Center; WRAP, Wisconsin Registry for Alzheimer’s Prevention.

### Clinical conversion

Parsimonious models for predicting conversion to AD and all-cause dementia are summarized in Table 3. Conversion to AD dementia was best predicted by CSF p-tau/Aβ42 ratio both in CU participants (BioFINDER-1 CU: AUC[95%CI]= 0.95 [0.93 – 0.97]) and MCI patients at baseline in the cohorts with data available (BioFINDER-1 MCI: AUC[95%CI]= 0.92 [0.89 – 0.95]; BioFINDER-2 MCI: AUC[95%CI]= 0.87[0.82 – 0.92]).

**Table 3.** Parsimonious model description for predicting clinical conversion. Description of the parsimonious model for predicting clinical conversion for each conversion type and cohort. These models were derived independently per each cohort and baseline diagnosis. Only those biomarkers selected to be into the parsimonious model per cohort and disease stage are shown in each case. Hazard ratios (HR) for each selected biomarker and AUC of the final model are shown. AUCs of the basic model (only covariates) are included for comparison. HRs were calculated with cox proportional hazards regression model with clinical conversion as the outcome. For all biomarkers, HRs represent increased risk of conversion for each SD change in biomarker value. To calculate AUC, we used generalized linear models including a binomial family. Covariates included in all models were: age, sex and *APOE-ε4* carriership. Abbreviations: Aβ, amyloid-β; AD, Alzheimer’s disease; AUC, area under the curve; CI, confidence interval; CU, cognitively unimpaired; HR, hazard ratio; MCI, mild cognitive impairment; NfL, neurofilament light; SD, standard deviation; p-tau; phosphorylated tau; WADRC, Wisconsin Alzheimer’s Disease Research Center; WRAP, Wisconsin Registry for Alzheimer’s Prevention.

| Conversion type | Cohort | Biomarker | HR<br>[95%CI] | p-value | AUC<br>[95%CI]<br>Parsimonious model | AUC<br>[95%CI]<br>basic model |
| --- | --- | --- | --- | --- | --- | --- |
| **CU →**<br>**AD dementia** | BioFINDER-1 | p-tau/aβ42 | 2.12<br>[1.80, 2.49] | <0.001 | 0.95<br>[0.93-0.97] | 0.78<br>[0.70-0.86] |
| **MCI →**<br>**AD dementia** | BioFINDER-1 | p-tau/aβ42 | 1.94<br>[1.68, 2.25] | <0.001 | 0.92<br>[0.89-0.95] | 0.77<br>[0.71-0.83] |
|  | BioFINDER-2 | p-tau/aβ42 | 2.35<br>[1.80, 3.08] | <0.001 | 0.87<br>[0.82-0.92] | 0.75<br>[0.68-0.82] |
| **CU →**<br>**All-cause**<br>**dementia** | BioFINDER-1 | p-tau/aβ42 | 1.65<br>[1.44, 1.90] | <0.001 | 0.90<br>[0.86-0.95] | 0.74<br>[0.67-0.80] |
|  |  | NfL | 2.09<br>[1.68, 2.61] | <0.001 |  |  |
| **MCI →**<br>**All-cause**<br>**dementia** | BioFINDER-1 | p-tau/aβ42 | 1.45<br>[1.26, 1.67] | <0.001 | 0.83<br>[0.77-0.89] | 0.71<br>[0.63-0.78] |
|  |  | NfL | 1.42<br>[1.23, 1.63] | <0.001 |  |  |
|  | BioFINDER-2 | aβ42/40 | 0.58<br>[0.42, 0.79] | <0.001 | 0.75<br>[0.68-0.82] | 0.62<br>[0.54-0.70] |
|  |  | NfL | 1.52<br>[1.25, 1.84] | <0.001 |  |  |
|  | WRAP &<br>WADRC | p-tau/aβ42 | 1.69<br>[1.05, 2.72] | 0.032 | 0.83<br>[0.72-0.94] | 0.73<br>[0.59-0.87] |

In the case of conversion to all-cause dementia, CSF NfL and CSF p-tau/Aβ42 ratio were included in the parsimonious model in CU participants at baseline (BioFINDER-1 CU: AUC[95%CI]= 0.90[0.86 – 0.95]). In participants that were MCI patients at baseline, in two out of three cohorts the CSF p-tau/Aβ42 ratio was included in the parsimonious model (BioFINDER-1 and WRAP & WADRC). In BioFINDER-1, CSF NfL was also included in the parsimonious model (BioFINDER-1 MCI: AUC[95%CI]= 0.83 [0.77 – 0.89]). In WRAP & WADRC, the CSF p-tau/Aβ42 ratio alone was sufficient to accurately predict conversion to all-cause dementia in MCI patients at baseline (AUC[95%CI]= 0.83[0.72 – 0.94]). In BioFINDER-2, the parsimonious model to predict conversion from MCI to all-cause dementia included the Aβ42/40 ratio and CSF NfL (AUC[95%CI]= 0.75[0.68 – 0.82]). However, this model was not significantly better than the one including the p-tau/Aβ42 ratio and NfL (AUC[95%CI]= 0.74[0.67 – 0.81], p=0.383).

We additionally calculated the HR for a one-SD increase in all CSF biomarkers selected in each of the parsimonious (Table 3). In summary, the CSF p-tau/Aβ42 ratio HRs [95% CI] for predicting conversion to AD dementia were 2.12 [1.80 – 2.49] in CU participants and around 2 in MCI patients at baseline (BioFINDER-1: HR[95%CI]= 1.94 [1.68 – 2.25]; BioFINDER-2: HR[95%CI]= 2.35 [1.84 – 3.08]). In the case of predicting conversion to all-cause dementia, HRs for the CSF p-tau/Aβ42 ratio were all around 1.5 both for CU participants (BioFINDER-1 CU: 1.65 [1.44 – 1.90]) and MCI patients at baseline (BioFINDER-1 MCI: 1.45 [1.23 – 1.63]; WRAP & WADRC MCI: 1.69[1.05 – 2.72]). HRs for NfL were all above 1.4 also in CU participants (BioFINDER-1 CU: 2.09 [1.68 – 2.61]) and MCI patients at baseline (BioFINDER-1 MCI: 1.42[1.23 – 1.63]; BioFINDER-2 MCI: 1.52[1.25 – 1.84]). Conversely, HR for the Aβ42/40 ratio (BioFINDER-2: 0.58[0.46 – 0.72]) was lower than 1 when predicting all-cause dementia from MCI. This was due to the well-known a negative association between the levels of this biomarker and pathology.

Receiver operating characteristic (ROC) curves for parsimonious, CSF p-tau/Aβ42 ratio-only and basic (*i*.*e*., only covariates) models are shown in Figure 2 and Kaplan-Meier curves for all the parsimonious models in Figure 3. Sensitivity analyses with conversion data truncated at 4- and 6-years follow-up can be found in Supplementary Tables 4 and 5, respectively. Sensitivity analyses where the p-tau/Aβ42 ratio was excluded are shown in Supplementary Table 6. All AUCs were equivalent or lower than models with the p-tau/Aβ42 ratio. To visualize this, we also included comparison plots between the individual markers and the p-tau/Aβ42 and Aβ42/40 ratios for BioFINDER-1 participants based on whether or not they converted to Alzheimer’s dementia (Supplementary Figure 2).

**Figure 2.**
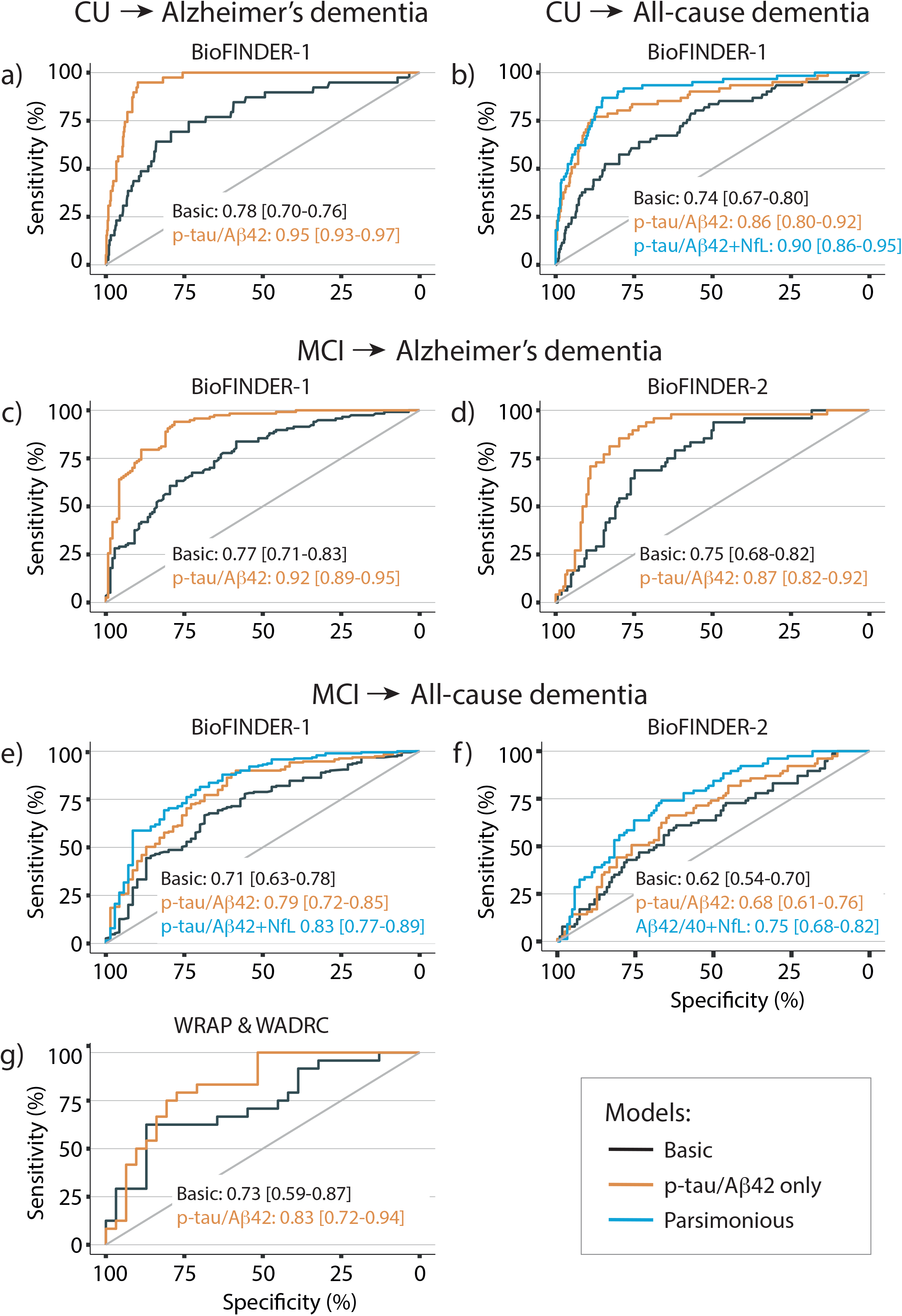
Clinical conversion. Depiction of ROC curves for basic, p-tau/aβ42 only and parsimonious models for predicting progression to AD or all-cause dementia. Models were created independently per clinical group at baseline (CU or MCI) and cohort (BioFINDER-1: [a], [c] and [e]; BioFINDER-2: [b], [d] and [f]; and WRAP & WADRC: [g]). Only scenarios with data available and more than 20 conversion cases are depicted. AUC[95%CI] are depicted for each model and case in the picture. Basic models only included covariates. Biomarkers included in the parsimonious models in each case are detailed in the figure. Covariates were included in all models and were: age, sex, *APOE-ε4* carriership and time. Abbreviations: Aβ, amyloid-β; AD, Alzheimer’s disease; AUC, area under the curve; CI, confidence interval; CU, cognitively unimpaired; MCI, mild cognitive impairment; NfL, neurofilament light; Ng, neurogranin; p-tau; phosphorylated tau; ROC, receiver operating characteristic; WADRC, Wisconsin Alzheimer’s disease Research Center; WRAP, Wisconsin Registry for Alzheimer’s Prevention.

**Figure 3.**
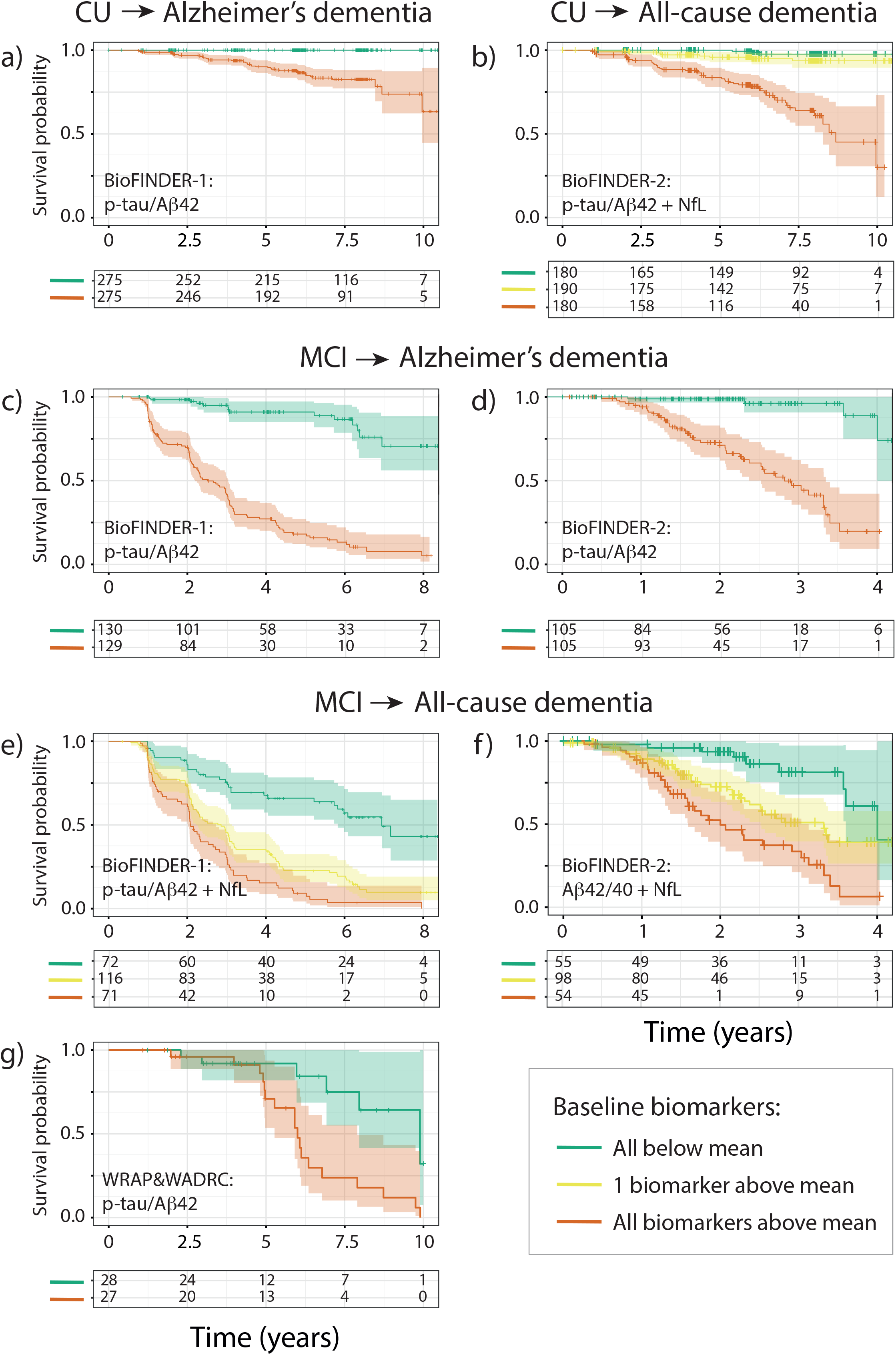
Kaplan-Meier survival curves for clinical conversion. Kaplan-Meier survival curves for each cohort and clinical group at baseline with conversion to Alzheimer’s dementia or all-cause dementia as outcome. Coloured lines depict different groups of individuals based on their baseline levels of selected biomarkers in the parsimonious models. Green (red) group line represents participants with all biomarkers in the parsimonious model below (above) the mean at baseline. For models with more than one biomarker included in the parsimonious model, yellow lines represent subjects with only one biomarkers above the mean at baseline. Coloured bands represent 95% confidence intervals and crosses represent censored data. These lines are only for visualization purposes and represent the mean trajectory of the group of participants included in these artificial groups. Individual trajectories were calculated per each participant in the statistical model. The total number of individuals in each group and timepoints are shown in the tables below the curves. Biomarkers used in the parsimonious model in each case are detailed in the plots. Abbreviations: Aβ, amyloid-β; CU, cognitively unimpaired; MCI, mild cognitive impairment; NfL, neurofilament light; Ng, neurogranin; p-tau; phosphorylated tau; WADRC, Wisconsin Alzheimer’s disease Research Center; WRAP, Wisconsin Registry for Alzheimer’s Prevention.

## Discussion

In this study, we have sought to identify the optimal combination of several CSF biomarkers analysed with recently developed fully automated assays for predicting disease progression in three large longitudinal cohorts. We found that the CSF p-tau/Aβ42 ratio alone or in combination with CSF NfL, a marker of neurodegeneration, may be sufficient to accurately predict disease progression both in cognitively unimpaired (CU) participants and in mild cognitive impairment (MCI) patients. The addition of CSF NfL may be especially important to predict conversion to all-cause dementia and, in some cases, cognitive decline. In contrast, other neurodegenerative markers and glial-related biomarkers did not significantly improve our predictive models. Further, we showed that the CSF p-tau/Aβ42 ratio was preferentially selected in the parsimonious models over the CSF Aβ42/40 ratio as a prognosis biomarker in AD. Although many previous studies have shown individual associations between some of these biomarkers and progression in AD and other dementias, this is the first study to investigate their prognosis utility as a combination of biomarkers and to compare it to typical core-AD biomarkers. Notably, our results have been replicated in three large independent cohorts with relatively long follow-up. Altogether, our study supports the use of the CSF p-tau/Aβ42 ratio, together with CSF NfL, in the clinical setting for prognosis of AD and other dementias.

The finding of CSF p-tau/Aβ42 ratio as the best marker for predicting disease progression was not unexpected. For many years now, the CSF p-tau/Aβ42 ratio has already shown high accuracy in predicting disease progression in both CU participants [34,35] and MCI patients [2,36]. The novelty of our study is that among a large panel of established and more novel CSF biomarkers, the CSF p-tau/Aβ42 ratio alone may be sufficient for an accurate disease prognosis, except for the addition of CSF NfL in some cases. Further, we also showed that the CSF p-tau/Aβ42 ratio may be more useful than the CSF Aβ42/40 ratio as a prognosis tool in AD. These results have important ramifications for clinical settings, where these biomarkers are increasingly available.

As aforementioned, we also found that CSF NfL was included in the parsimonious models for predicting disease progression in particular scenarios. In this study, higher levels of CSF NfL have consistently reported higher hazard ratios for converting to *all-cause* dementia, both for CU and MCI groups. In line with our results, previous studies have already shown the tight association between CSF NfL and brain and cognitive deterioration [37]. The fact that CSF NfL improved the prediction models of conversion to *all-cause* dementia but not to *Alzheimer’s* dementia was in line with previous findings of CSF NfL as a non-specific biomarker of neurodegeneration [5,38]. Thus, based on our results, CSF NfL may be helpful to predict conversion to dementias other than Alzheimer’s, which in our models was already sufficiently captured by the CSF p-tau/Aβ42 ratio.

We found that the addition of CSF NfL to the CSF p-tau/Aβ42 ratio, was also useful for predicting cognitive decline. This was only significant in the case of mPACC change prediction in BioFINDER-1 (n=550). However, this CU group had a longer follow-up than in BioFINDER-2 (BioFINDER-1 mean follow-up time: 6.2 years *vs*. BioFINDER-2 mean follow-up time: 1.2 years) and was significantly older than in the joint WRAP & WADRC group (BioFINDER-1 mean age: 71.8 years old *vs*. WRAP & WADRC mean age: 61.2 years old). These characteristics may have facilitated the detection of a larger change in cognition, which was not only attributed to CSF p-tau/Aβ42 ratio. Further supporting the utility of CSF NfL for predicting cognitive decline, it is important to note that it was the most frequently selected biomarker, after the CSF p-tau/Aβ42 ratio, although in most cases its addition did not significantly improve the model. Nonetheless, we cannot disregard the possibility of a false positive result for CSF NfL. Future studies may benefit from including a longer follow-up and a more diverse population to elucidate the full clinical value of CSF NfL.

In recent years, there has been increasing interest in investigating glial activation markers and their relationship with disease progression, as it has been suggested that inflammatory processes may have an active role in AD [4]. Previous studies proposed that some glial activation markers could be related to disease progression [7,8]. For instance, higher levels of sTREM2, a microglial activation marker [39], have been related to slower rates of Aβ accumulation and disease progression [40–42]. Levels of YKL-40, an astrocytic marker, were also proposed as a potential prognostic marker of AD [43]. However, those studies were conducted without their comparison to core AD biomarkers or if so, they did not show a significant improvement on their prediction accuracy. Similarly, neurodegenerative markers other than CSF NfL were not included in our final models. Altogether, our results suggest that, although their study hold great importance to understand the biological processes underlying the course of AD, glial activation markers and neurodegeneration markers other than NfL do not provide additional value in predicting disease progression to justify their use in a clinical setting. The main strength of this study is that analyses were performed in three large longitudinal cohorts. Although there were small differences in particular models, parsimonious models for each condition were replicated in all cohorts, reinforcing our results. Furthermore, we used a set of CSF biomarkers measured with the same technique and in the same single panel, which reduces variability in our measures. Nonetheless, some limitations must be acknowledged. First, we focused on CSF biomarkers rather than the recently developed plasma biomarkers. Plasma biomarkers are promising but are, in contrast with CSF, not yet widely available in clinical practice [12,44–47]. Second, participants were recruited from cohort studies, which may limit the generalizability of our results to a more representative clinical population. Nonetheless, BioFINDER-2 is a study not only focused on AD and include participants with all types of dementias; and BioFINDER-1 is a representative sample of patients from a Memory Clinic in Sweden, which increases the diversity of the sample. Third, although our mean of follow-up time was considerably long, it may still not be sufficient to capture cognitive decline in CU, as most neurodegenerative disorders show a relatively gradual decline. To alleviate this caveat, we used the mPACC to measure cognitive decline in CU, which is specifically developed to detect the first signs of cognitive decline in a cognitively normal population [29]. Nonetheless, results of clinical progression in this group may be biased due to the low number of conversions, specially to Alzheimer’s dementia, and for this reason our results should be considered with caution. Finally, we acknowledge that although the p-tau/Aβ42 ratio showed prognostic utility, it may not be the most adequate biomarker to assess the actual brain pathology.

Our results suggest that the CSF p-tau/Aβ42 ratio is sufficient, compared with other CSF biomarkers, to accurately predict disease progression in three large longitudinal cohorts. The addition of CSF NfL to the CSF p-tau/Aβ42 ratio may improve prediction of progression to all-cause dementia and cognitive decline in some cases. On the other hand, other markers of neurodegeneration and glial activation markers do not seem to provide additional value on disease progression prediction. These results mays be useful for its implementation to the clinical setting although further research in more diverse populations is needed.

## Data Availability

All data produced in the present study are available upon reasonable request to the authors

## Acknowledgements

Work at the authors’ research center was supported by the Swedish Research Council (2016-00906), the Knut and Alice Wallenberg foundation (2017-0383), the Marianne and Marcus Wallenberg foundation (2015.0125), the Strategic Research Area MultiPark (Multidisciplinary Research in Parkinson’s disease) at Lund University, the Swedish Alzheimer Foundation (AF-939932), the Swedish Brain Foundation (FO2021-0293), The Parkinson foundation of Sweden (1280/20), the Konung Gustaf V:s och Drottning Victorias Frimurarestiftelse, the Skåne University Hospital Foundation (2020-O000028), Regionalt Forskningsstöd (2020-0314) and the Swedish federal government under the ALF agreement (2018-Projekt0279). SP is supported by Swedish Research Council (2018-02052), the Swedish Brain Foundation (FO2020-0271) and the Swedish Alzheimer Foundation (AF-940046). HZ is a Wallenberg Scholar supported by grants from the Swedish Research Council (#2018-02532), the European Research Council (#681712 and #101053962), Swedish State Support for Clinical Research (#ALFGBG-71320), the Alzheimer Drug Discovery Foundation (ADDF), USA (#201809-2016862), the AD Strategic Fund and the Alzheimer’s Association (#ADSF-21-831376-C, #ADSF-21-831381-C and #ADSF-21-831377-C), the Olav Thon Foundation, the Erling-Persson Family Foundation, Stiftelsen för Gamla Tjänarinnor, Hjärnfonden, Sweden (#FO2019-0228), the European Union’s Horizon 2020 research and innovation programme under the Marie Skłodowska-Curie grant agreement No 860197 (MIRIADE), the European Union Joint Programme – Neurodegenerative Disease Research (JPND2021-00694), and the UK Dementia Research Institute at UCL (UKDRI-1003). KB is supported by the Swedish Research Council (#2017-00915), the Alzheimer Drug Discovery Foundation (ADDF), USA (#RDAPB-201809-2016615), the Swedish Alzheimer Foundation (#AF-742881), Hjärnfonden, Sweden (#FO2017-0243), the Swedish state under the agreement between the Swedish government and the County Councils, the ALF-agreement (#ALFGBG-715986), and European Union Joint Program for Neurodegenerative Disorders (JPND2019-466-236). The University of Wisconsin authors (KAC, EMJ, SCJ) and data were supported by NIH (R01AG027161, RO1AG021155, P30AG062715), and the University of Wisconsin Institute for Clinical and Translational Research NCATS (TL1TR002375).

The funding sources had no role in the design and conduct of the study; in the collection, analysis, interpretation of the data; or in the preparation, review, or approval of the manuscript.

