## Supplementary Material for "Optimal combinations of CSF biomarkers for predicting cognitive decline and clinical conversion in cognitively unimpaired participants and mild cognitive impairment patients: A multi-cohort study"

**Supplementary Table 1. Parsimonious model description for predicting cognitive decline at 4-years follow-up**

| Diagnosis group | Cohort | Biomarker | β [95% CI] | BIC  parsimonious | R^2^ parsimonious | BIC  basic | R^2^ basic |
| --- | --- | --- | --- | --- | --- | --- | --- |
| CU | BioFINDER-1 | p-tau/Aβ42 | -0.09  [-0.12, -0.05] | 2651.6 | 0.24 | 2731.1 | 0.08 |
|  |  | NfL | -0.07  [-0.11, -0.04] |  |  |  |  |
|  | BioFINDER-2 | p-tau/Aβ42 | -0.10  [-0.14, -0.05] | 1297.9 | 0.36 | 1314.1 | 0.27 |
|  | WRAP & WADRC | p-tau/Aβ42 | -0.09  [-0.12, -0.06] | 2686.3 | 0.32 | 2708.6 | 0.29 |
| MCI | BioFINDER-1 | p-tau/Aβ42 | -0.18  [-0.24, -0.13] | 2207.9 | 0.31 | 2234.4 | 0.22 |
|  | BioFINDER-2 | p-tau/Aβ42 | -0.15  [-0.24, -0.06] | 976.6 | 0.19 | 978.8 | 0.13 |
|  | WRAP & WADRC | p-tau/Aβ42 | -0.25  [-0.37, -0.13] | 475.0 | 0.37 | 488.0 | 0.13 |

Description of the parsimonious model for predicting cognitive decline for each diagnosis group and cohort including effect sizes (β) for each selected biomarker and statistics (BIC and R^2^) of the final model. Only data collected up to 4-years (+6 months) follow-up was included in this analysis. Linear mixed models with random time slope and intercept were used in all cases. β estimates represent the effect size of each biomarker’s interaction with time. Cognitive decline was assessed with mPACC in CU and with MMSE in MCI patients. Covariates were included in all models and were: age, sex, *APOE-ε4* carriership, education and time.

Abbreviations: Aβ, amyloid-β; BIC, Bayesian Information Criterion; CI, confidence interval; CU, cognitively unimpaired; MCI, mild cognitive impairment; MMSE, Mini-Mental State Examination; mPACC, modified Preclinical Alzheimer Cognitive Composite; p-tau, phosphorylated tau; REML, Restricted Maximum Likelihood; WADRC, Wisconsin Alzheimer’s disease Research Center; WRAP, Wisconsin Registry for Alzheimer’s Prevention.

**Supplementary Table 2. Parsimonious model description for predicting cognitive decline at 6-years follow-up**

| Diagnosis group | Cohort | Biomarker | β [95% CI] | BIC  parsimonious | R^2^ parsimonious | BIC  basic | R^2^ basic |
| --- | --- | --- | --- | --- | --- | --- | --- |
| CU | BioFINDER-1 | p-tau/Aβ42 | -0.15  [-0.19, -0.12] | 3110.8 | 0.25 | 3231.6 | 0.06 |
|  |  | NfL | -0.08  [-0.12, -0.05] |  |  |  |  |
|  | WRAP & WADRC | p-tau/Aβ42 | -0.11  [-0.13, -0.08] | 3244.2 | 0.31 | 3287.7 | 0.27 |
| MCI | BioFINDER-1 | p-tau/Aβ42 | -0.22  [-0.28, -0.16] | 2364.1 | 0.39 | 2397.7 | 0.28 |
|  | WRAP & WADRC | p-tau/Aβ42 | -0.27  [-0.41, -0.13] | 531.7 | 0.37 | 544.5 | 0.13 |

Description of the parsimonious model for predicting cognitive decline for each diagnosis group and cohort including effect sizes (β) for each selected biomarker and statistics (BIC and R^2^) of the final model. Only data collected up to 6-years (+6 months) follow-up was included in this analysis. Linear mixed models with random slope and intercept were used in all cases. β estimates represent the effect size of each biomarker’s interaction with time. Cognitive decline was assessed with mPACC in CU and with MMSE in MCI patients. Covariates were included in all models and were: age, sex, *APOE-ε4* carriership, education and time. BioFINDER-2 cohort was not included in this table as the maximum follow-up was at 4-years after baseline.

Abbreviations: Aβ, amyloid-β; BIC, Bayesian Information Criterion; CI, confidence interval; CU, cognitively unimpaired; MCI, mild cognitive impairment; MMSE, Mini-Mental State Examination; mPACC, modified Preclinical Alzheimer Cognitive Composite; p-tau, phosphorylated tau; REML, Restricted Maximum Likelihood; WADRC, Wisconsin Alzheimer’s disease Research Center; WRAP, Wisconsin Registry for Alzheimer’s Prevention.

**Supplementary Table 3. Parsimonious model description for predicting clinical conversion at 4-years follow-up**

| Conversion type | Cohort | Biomarker | HR  [95%CI] | p-value | AUC [95%CI]  Parsimonious model | AUC [95%CI] basic model |
| --- | --- | --- | --- | --- | --- | --- |
| CU → AD dementia | BioFINDER-1 (n=21) | p-tau/aβ42 | 2.14  [1.77, 1.58] | <0.001 | 0.96  [0.94-0.98] | 0.73  [0.61-0.86] |
| MCI → AD dementia | BioFINDER-1 (n=102) | p-tau/aβ42 | 1.98  [1.71, 2.31] | <0.001 | 0.91  [0.88-0.95] | 0.72  [0.66-0.78] |
|  | BioFINDER-2 (n=50) | p-tau/aβ42 | 2.35  [1.80, 3.08] | <0.001 | 0.87  [0.82-0.92] | 0.74  [0.66-0.81] |
| CU → All-cause dementia | BioFINDER-1 (n=31) | p-tau/aβ42 | 1.62  [1.37, 1.91] | <0.001 | 0.92  [0.88-0.95] | 0.70  [0.60-0.80] |
|  |  | NfL | 2.21  [1.66, 2.95] | <0.001 |  |  |
| MCI → All-cause dementia | BioFINDER-1 (n=166) | p-tau/aβ42 | 1.45  [1.25, 1.68] | <0.001 | 0.79  [0.73-0.85] | 0.66  [0.59-0.73] |
|  |  | NfL | 1.39  [1.20, 1.61] | <0.001 |  |  |
|  | BioFINDER-2 (n=77) | aβ42/40 | 0.60  [0.44, 0.82] | 0.002 | 0.75  [0.68-0.82] | 0.62  [0.54-0.70] |
|  |  | NfL | 1.48  [1.22, 1.79] | <0.001 |  |  |
|  | WRAP&WADRC (n=21) | NfL | 1.24  [0.89, 1.72] | 0.202 | 0.84  [0.74-0.95] | 0.75  [0.62 – 0.89] |

Description of the parsimonious model for predicting clinical conversion for each conversion type and cohort including hazard ratios (HR) for each selected biomarker and AUC of the final model at 4-years (+6 months) follow-up. If participants converted after 4-years follow-up, then they were considered non-converters. Hazard ratios were calculated with cox proportional hazards regression model with clinical conversion as outcome. For all biomarkers, hazard ratios represent increased risk of conversion for each SD change in biomarker value. To calculate AUC, we used generalized linear models including a binomial family. Covariates included in all models were: age, sex and *APOE-ε4* carriership. Number of conversions for each cohort and conversion type can be found after each cohort’s name.

Abbreviations: Aβ, amyloid-β; AD, Alzheimer’s disease; AUC, area under the curve; CI, confidence interval; CU, cognitively unimpaired; HR, hazard ratio; MCI, mild cognitive impairment; NfL, neurofilament light; SD, standard deviation; p-tau; phosphorylated tau; WADRC, Wisconsin Alzheimer’s disease Research Center; WRAP, Wisconsin Registry for Alzheimer's Prevention.

**Supplementary Table 4. Parsimonious model description for predicting clinical conversion at 6-years follow-up**

| Conversion type | Cohort | Biomarker | HR  [95%CI] | p-value | AUC [95%CI]  Parsimonious model | AUC [95%CI] basic model |
| --- | --- | --- | --- | --- | --- | --- |
| CU →  AD dementia | BioFINDER-1 (n=35) | p-tau/aβ42 | 2.12  [1.80, 2.50] | <0.001 | 0.95  [0.93-0.97] | 0.77  [0.68-0.86] |
| MCI → AD dementia | BioFINDER-1 (n=114) | p-tau/aβ42 | 2.88  [2.33, 3.57] | <0.001 | 0.91  [0.88-0.95] | 0.76  [0.70-0.82] |
| CU → All-cause dementia | BioFINDER-1 (n=49) | p-tau/aβ42 | 1.90  [1.65, 2.18] | <0.001 | 0.86  [0.79-0.92] | 0.72  [0.64-0.80] |
| MCI → All-cause dementia | BioFINDER-1 (n=185) | p-tau/aβ42 | 1.45  [1.25, 1.67] | <0.001 | 0.82  [0.76-0.88] | 0.69  [0.61-0.76] |
|  |  | NfL | 1.43  [1.24, 1.65] | <0.001 |  |  |
|  | WRAP&WADRC (n=22) | NfL | 1.58  [1.12, 2.24] | 0.009 | 0.85  [0.76-0.95] | 0.74  [0.60 – 0.88] |

Description of the parsimonious model for predicting clinical conversion for each conversion type and cohort including hazard ratios (HR) for each selected biomarker and AUC of the final model at 6-years (+6 months) follow-up. If participants converted after 6-years follow-up, then they were considered non-converters. HRs were calculated with cox proportional hazards regression model with clinical conversion as outcome. For all biomarkers, hazard ratios represent increased risk of conversion for each SD change in biomarker value. To calculate AUC, we used generalized linear models including a binomial family. Covariates included in all models were: age, sex and *APOE-ε4* carriership. Number of conversions for each cohort and conversion type can be found after each cohort’s name. BioFINDER-2 cohort was not included in this table as the maximum follow-up was at 4-years after baseline.

Abbreviations: Aβ, amyloid-β; AD, Alzheimer’s disease; AUC, area under the curve; CI, confidence interval; CU, cognitively unimpaired; HR, hazard ratio; MCI, mild cognitive impairment; NfL, neurofilament light; SD, standard deviation; p-tau; phosphorylated tau; WADRC, Wisconsin Alzheimer’s disease Research Center; WRAP, Wisconsin Registry for Alzheimer's Prevention.

**Supplementary Table 5. Parsimonious model description for predicting clinical conversion without including the p-tau/Aβ42 ratio**

| Conversion type | Cohort | Biomarker | HR  [95%CI] | p-value | AUC [95%CI]  Parsimonious model | AUC [95%CI] basic model |
| --- | --- | --- | --- | --- | --- | --- |
| CU $\boldsymbol{\to}$  AD dementia | BioFINDER-1 | Aβ42/40 | 0.13  [0.07, 0.24] | <0.001 | 0.93  [0.90-0.95] | 0.78  [0.70-0.86] |
| MCI $\boldsymbol{\to}$  AD dementia | BioFINDER-1 | Aβ42/40 | 0.30  [0.20, 0.44] | <0.001 | 0.92  [0.89-0.95] | 0.77  [0.71-0.83] |
|  |  | p-tau | 1.25  [1.02, 1.53] | 0.031 |  |  |
|  | BioFINDER-2 | Aβ42/40 | 0.26  [0.13, 0.51] | <0.001 | 0.88  [0.83-0.92] | 0.75  [0.68-0.82] |
|  |  | p-tau | 1.37  [0.99, 1.89] | 0.056 |  |  |
| CU $\boldsymbol{\to}$  All-cause dementia | BioFINDER-1 | Aβ42 | 0.46  [0.33, 0.63] | <0.001 | 0.89  [0.85-0.94] | 0.74  [0.67-0.80] |
|  |  | NfL | 2.10  [1.73, 2.55] | <0.001 |  |  |
| MCI $\boldsymbol{\to}$  All-cause dementia | BioFINDER-1 | Aβ42 | 0.59  [0.49, 0.70] | <0.001 | 0.84  [0.78-0.90] | 0.71  [0.63-0.78] |
|  |  | NfL | 1.46  [1.28, 1.67] | <0.001 |  |  |
|  | BioFINDER-2 | Aβ42 | 0.53  [0.37, 0.76] | <0.001 | 0.74  [0.67-0.81] | 0.62  [0.54-0.70] |
|  |  | NfL | 1.55  [1.28, 1.87] | <0.001 |  |  |
|  | WRAP & WADRC | - | - | - | - | 0.73  [0.59-0.87] |

Description of the parsimonious model for predicting clinical conversion for each conversion type and cohort including hazard ratios (HR) for each selected biomarker and AUC of the final model. HRs were calculated with cox proportional hazards regression model with clinical conversion as outcome. For all biomarkers, hazard ratios represent increased risk of conversion for each SD change in biomarker value. To calculate AUC, we used generalized linear models including a binomial family. Covariates included in all models were: age, sex and *APOE-ε4* carriership.

Abbreviations: Aβ, amyloid-β; AD, Alzheimer’s disease; AUC, area under the curve; CI, confidence interval; CU, cognitively unimpaired; HR, hazard ratio; MCI, mild cognitive impairment; NfL, neurofilament light; SD, standard deviation; p-tau; phosphorylated tau; WADRC, Wisconsin Alzheimer’s disease Research Center; WRAP, Wisconsin Registry for Alzheimer's Prevention.

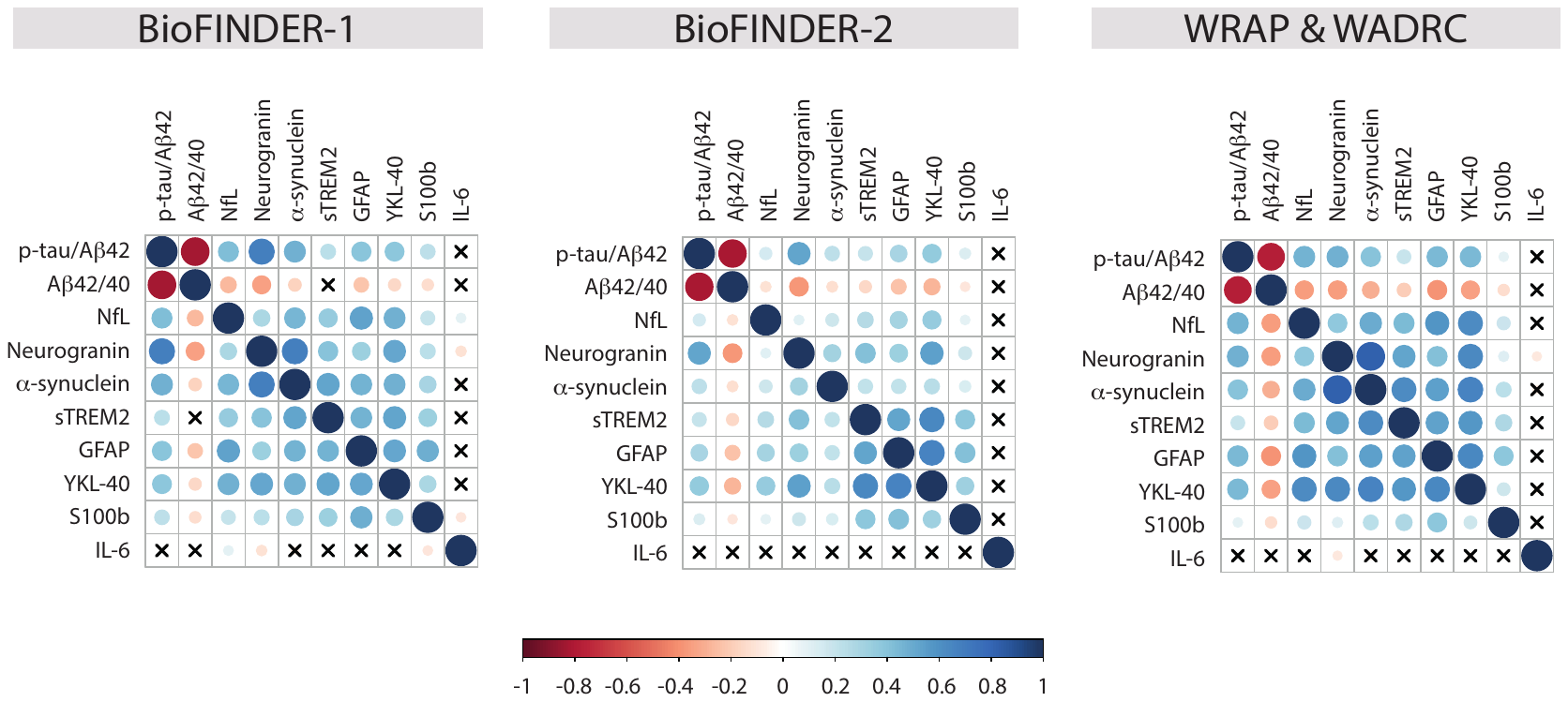

**Supplementary Figure 1. Biomarkers’ cross-correlation**

Correlation among biomarkers calculated by Spearman’s ρ. Circle size corresponds to the effect size. Blue (red) circles represent positive (negative) associations. Crosses represent non-significant (p>0.05) associations. CU participants and MCI patients have been included in this analysis.

Abbreviations: Aβ, amyloid-β; CU, cognitively unimpaired; GFAP, glial fibrillary acidic protein; IL-6, interleukin 6; MCI, mild cognitive impairment; NfL, neurofilament light; p-tau; phosphorylated tau; sTREM2, soluble triggering receptor expressed on myeloid cells 2; S100b, S100 calcium-binding protein B; YKL-40, chitinase 3-like 1; WADRC, Wisconsin Alzheimer’s disease Research Center; WRAP, Wisconsin Registry for Alzheimer's Prevention.

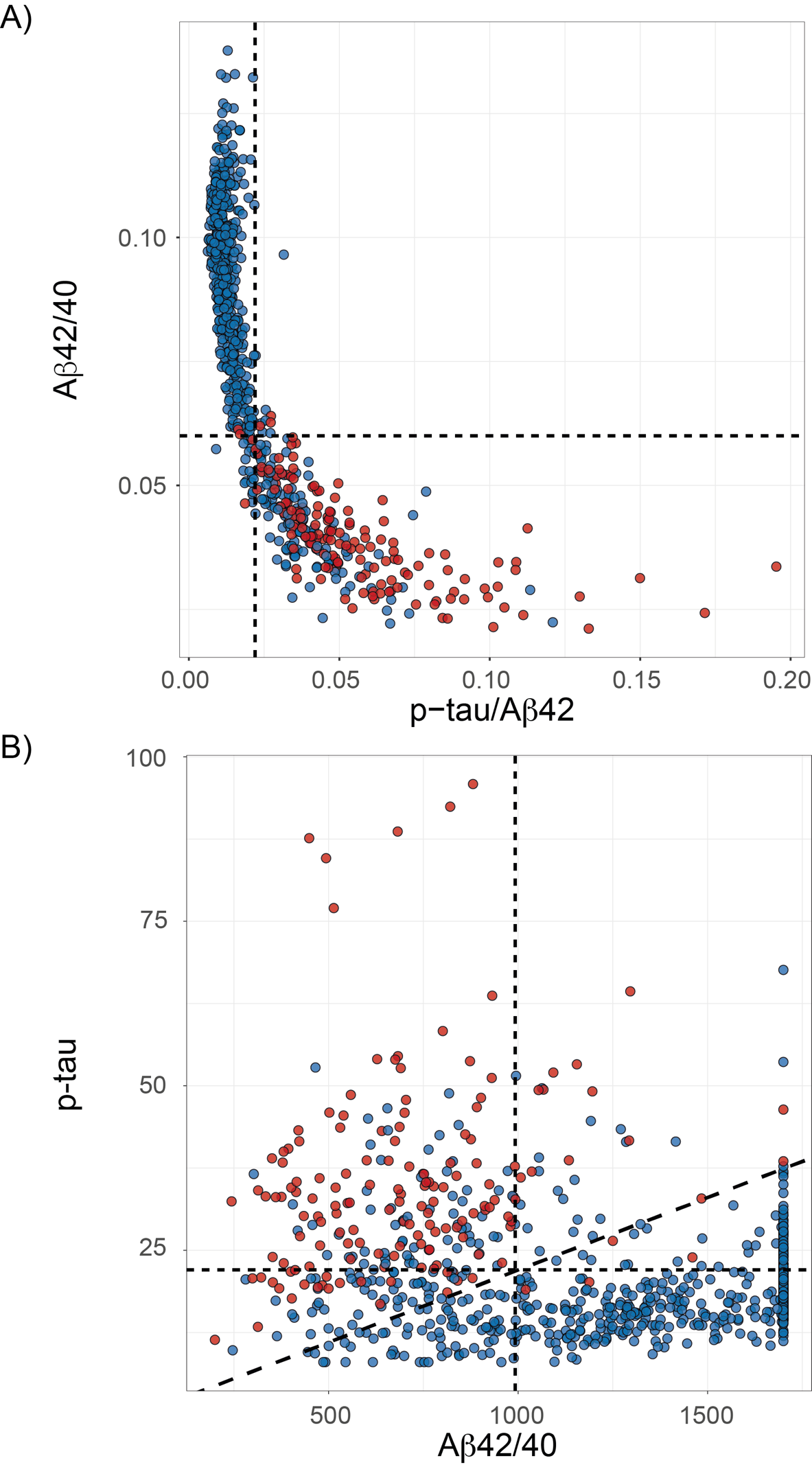

**Supplementary Figure 2. Individual vs ratio biomarkers for predicting conversion to Alzheimer’s dementia in BioFINDER-1**

Association between CSF Aβ42/40 ratio and p-tau/Aβ42 ratio (A) and between CSF p-tau and Aβ42 (B) in the whole BioFINDER-1 cohort. Red dots represent participants who converted to Alzheimer’s dementia in the follow-up and blue, those that did not. Dashed lines represent optimal cut-offs for predicting conversion to Alzheimer’s dementia for each biomarker (A: vertical p-tau/Aβ42=0.022 and horizontal Aβ42/40=0.060; B: vertical Aβ42: 992.7, horizontal p-tau: 21.99, diagonal p-tau/Aβ42: 0.022).

Abbreviations: Aβ, amyloid-β.
